# Preindex Social Context and Cardiovascular Events Across Blood Pressure, LDL Cholesterol, and A1c Cohorts

**DOI:** 10.64898/2026.09.28.26364240

**Authors:** Sana Mirza, Joshua Moen

**Affiliations:** Physician Assistant Program, UC Davis Health, Sacramento, California, USA

**Author notes:** Correspondence: Joshua Moen, PhD, MPH, MSPAS, Physician Assistant Program, UC Davis Health, 4301 X Street, Sacramento, CA, USA Correspondence email: Joshua Moen. Author emails: Sana Mirza.

**Keywords:** cardiovascular events, social determinants of health, blood pressure, LDL cholesterol, hemoglobin A1c, All of Us

## Abstract

**Background:** Cardiovascular prevention is organized around target metrics. Blood pressure, LDL cholesterol, and A1c define treatment targets, quality metrics, and, implicitly, who is considered safe. Yet a value below a cutoff is a correlative measurement, not an outcome. We tested whether social context recorded before the measurement separated subsequent cardiovascular events among people whose numbers already met the cutoff.

**Methods:** In this retrospective cohort study, we analyzed All of Us participants without recorded cardiovascular disease and with low comorbidity in three separately indexed cohorts: systolic blood pressure (SBP), LDL cholesterol, and A1c. A preindex score combined household income, education, food security, physical activity, and ZIP-linked deprivation, standardized against a fixed pooled reference of 325,652 eligible people. Cox models adjusted for the measured biomarker value, its square, a score-by-biomarker term, medication exposure, and clinical covariates. The outcome was first myocardial infarction, ischemic stroke, or heart failure. A sensitivity analysis indexed participants at the first measurement after the score became computable and counted only events recorded in inpatient or emergency settings.

**Results:** The SBP, LDL, and A1c cohorts included 19,525, 16,578, and 18,539 participants with 144, 129, and 191 events. Each one-SD more favorable score was associated with lower hazard: HR 0.611 (95% CI 0.498–0.749), 0.631 (0.512–0.778), and 0.544 (0.457–0.648), respectively. Below each study cutoff (SBP <140 mmHg, LDL <100 mg/dL, A1c <8%), less favorable context had event rates of 3.39 versus 1.68, 7.16 versus 2.05, and 8.67 versus 2.34 per 1,000 person-years. In a one-year analysis restricted to 2019–2023 index dates, the corresponding adjusted HRs were 0.530, 0.336, and 0.365. In the sensitivity cohorts (54,338, 41,451, and 33,607 participants; 493, 288, and 316 events), HRs were 0.732 (0.657–0.815), 0.660 (0.573–0.761), and 0.631 (0.552–0.722).

**Conclusions:** A biomarker below the study cutoff did not identify a uniform, low-risk population. Social context recorded before the measurement separated cardiovascular events after adjustment for the measured value and clinical factors. Risk assessment that stops at the number misses this variation.

**What Is Known:**

- Social, economic, and neighborhood conditions shape cardiovascular risk factors and predict cardiovascular events.
- Blood pressure, LDL cholesterol, and A1c targets anchor prevention and quality measurement, yet trials of treating to those targets show modest absolute reductions in hard outcomes.

**What the Study Adds:**

- After adjustment for the measured biomarker itself, each SD of more favorable preindex social context was associated with a 37% to 46% lower hazard of myocardial infarction, ischemic stroke, or heart failure across SBP, LDL cholesterol, and A1c cohorts. The association persisted, at 27% to 37%, in sensitivity cohorts 1.8 to 2.8 times larger.
- Below each biomarker cutoff, less favorable context carried 2- to nearly 4-fold higher observed event rates.
- A value below a study cutoff identifies a biomarker category; recorded social context identifies variation in subsequent events within that category.

## Introduction

Cardiovascular prevention runs on numbers; blood pressure, LDL cholesterol, and A1c are often measured quarterly to annually, anchor guideline targets, and serve as performance measures for clinicians and health systems (1–4). The reasoning is straightforward: move the number below a line, and expect risk to move with it. Yet the number is a proxy while the outcomes patients experience, myocardial infarction, stroke, and heart failure, are what cardiovascular studies ultimately count (5). If people with similarly favorable values go on to have different event rates, the value alone does not describe their subsequent risk.

Social context is the obvious place to look for what the number leaves out. Powell-Wiley and colleagues describe a pathway that runs from structural conditions through daily social and environmental exposures to biological responses, conventional risk factors, and cardiovascular outcomes (6). In that framework, a clinic measurement is a downstream snapshot of a longer process, not a complete account of it. Individual- and neighborhood-level social conditions predict subsequent cardiovascular events (7–9), yet these associations have seldom been examined inside the biomarker categories that actually drive clinical decisions.

Cardiology now has standardized data elements for social determinants (10), All of Us supports a survey-based deprivation index (11), and multidomain cardiovascular health scores predict incident disease (12). Neighborhood vulnerability tracks with midlife cardiovascular health (13) and with referral to cardiac rehabilitation (14), and risk equations such as PREVENT estimate who will develop disease (15). What remains unanswered is the question most relevant to target-based care: among people whose measured values already fall below the same cutoff, does social context identify who will have an event?

We addressed that question in three cohorts indexed separately on systolic blood pressure (SBP), LDL cholesterol, and A1c. We calculated available social-context scores against one fixed reference, then compared subsequent myocardial infarction, ischemic stroke, or heart failure after accounting for each person’s actual biomarker value and recorded clinical characteristics. We also examined the four combinations of context and biomarker category. The analysis asks how much variation in clinical outcomes remains within a favorable measurement category and whether recorded social context identifies it.

## Methods

### Data source and cohorts

We used the All of Us Researcher Workbench Controlled Tier curated data repository C2025Q4R6. Separate prevalent-measurement cohorts were indexed on the first qualifying SBP, LDL cholesterol, or A1c measurement at least 365 days after the first recorded observation. Eligibility required no recorded prior cardiovascular disease and an age-adjusted Charlson score (16) of 2 or less. The SBP, LDL, and A1c eligible source cohorts contained 306,353, 161,937, and 126,504 participants, respectively. A person could enter more than one biomarker cohort; each cohort was modeled separately. The final analysis required a computable context score, preindex estimated glomerular filtration rate (eGFR) of at least 60 mL/min/1.73 m^2^, a measured index biomarker, and positive follow-up time. Eligible index dates spanned 1986-02-18 to 2025-01-01 for SBP, 1986-01-21 to 2025-01-01 for LDL, and 1986-02-07 to 2025-01-01 for A1c. Analyzed index dates spanned 2017-06-06 to 2024-12-30, 2017-06-30 to 2024-12-31, and 2017-07-22 to 2025-01-01, respectively. In the eligible cohorts, recorded observation end dates spanned 2009-12-01 to 2025-01-02, 2013-09-25 to 2025-01-02, and 2013-12-16 to 2025-01-02, respectively.

### Preindex social context score

We summarized recorded context with five components: ordinal household income, years of education, food security, activity, and ZIP-linked deprivation. Activity combined walking and moderate-activity responses. For each survey item, we used the latest valid answer recorded on or before the biomarker index date; the geographic deprivation value came from the linked area-level source data. This timing placed the survey information before the outcomes being studied.

To make the score comparable across SBP, LDL, and A1c cohorts, we estimated each component’s mean and standard deviation in one pooled reference of 325,652 unique eligible people. The earliest eligible index supplied reference values for people entering more than one cohort. We standardized each component against that reference. A person needed at least three observed components; we summed those available and multiplied by five divided by the number observed, preserving a five-component scale. The reference score was computable for 65,374 people. Its median, −0.2573, defined the more- and less-favorable categories in descriptive analyses, and its standard deviation, 3.5723, defined the unit of the continuous-score hazard ratios. These reference values and the cutoff stayed fixed across cohorts.

### Outcome and covariates

The outcome was the first recorded postindex myocardial infarction, ischemic stroke, or heart failure. Follow-up ended at the first event or the recorded observation end. The earliest qualifying postindex event date defined MACE timing. The measured index SBP, LDL, or A1c value entered its model directly. Drug eras were classified as active at index or ended within the preceding 180 days. Additional covariates were age, sex, Black and Hispanic indicators, the age-adjusted Charlson score, preindex eGFR, preindex smoking and its missingness flag, prior-year visit count, and index year. The eGFR calculation used the 2021 CKD-EPI creatinine equation with the closest creatinine result in the preceding 365 days (17). An absent preindex smoking response was represented by the smoking-missingness indicator. Participants without a computable context score or preindex eGFR were excluded from the primary models.

### Statistical analysis

We fit separate unpenalized Cox models for each biomarker cohort. Each model included the continuous social-context score per one pooled-reference standard deviation, the observed biomarker value standardized within its analysis cohort, its squared term, and a score-by-biomarker term, along with the covariates listed above. The reported score hazard ratio is evaluated at the cohort mean biomarker value. Ninety-five percent confidence intervals used the model-based Cox variance. We also tabulated observed event counts and person-time across the four combinations of score category and measured biomarker category. The fixed descriptive cutoffs were SBP below 140 mmHg, LDL below 100 mg/dL, and A1c below 8%. All four strata were calculated in each cohort. All participants remained in the cohort totals and Cox models; All of Us disclosure rules affected only which descriptive rows and counts could be displayed in Table 3. These cutoffs define the study categories; clinical treatment goals can vary by patient.

The restricted-era sensitivity retained index dates in 2019–2023, censored follow-up at one year, and used a separate baseline hazard for each index year. Its focused adjustment set comprised the context score, measured biomarker and its square, age, sex, age-adjusted Charlson score, and preindex eGFR. Participants censored before one year remained in the risk sets until their recorded follow-up ended. We reported one-year event counts and the number with a one-year event or at least one year of observation. Participant flow, component availability, and differences between analyzed and other eligible participants were audited in the corrected source cohorts, consistent with cohort reporting guidance (18).

The primary design fixed the index at the first qualifying measurement and then required a score computable from answers recorded on or before that date. Because most participants answered the surveys after their first qualifying measurement, this design retained 6.4% to 14.7% of eligible participants. To test whether that selection explained the findings, we repeated the analysis with the index moved to the first qualifying measurement on or after the date the score became computable, defined as the date the third of the five components was recorded. Eligibility, the preindex timing of survey answers and creatinine, the eGFR requirement, and the three-of-five rule were otherwise unchanged. Component means and standard deviations were re-estimated with the same fixed-reference method among unique eligible people in these cohorts. In this analysis, myocardial infarction and ischemic stroke counted only when recorded at an inpatient, emergency department, or emergency-to-inpatient visit, and heart failure only at an inpatient or emergency-to-inpatient visit. Cox models included the score per reference standard deviation, the measured biomarker standardized within cohort and its square, medication exposure, and the primary covariates. They also included age squared, insurance status with a missingness indicator, family history of cardiovascular disease, and emergency department visits in the prior year.

Biomarker values were winsorized at the 0.5th and 99.5th percentiles, and the score-by-biomarker term was tested separately by likelihood ratio. We also refit these models retaining participants without a preindex eGFR, using a missingness indicator, and repeated the four-group rate tabulation and the 2019–2023 one-year analysis, adding medication exposure to the focused adjustment set.

## Results

### REFERENCE score and analysis populations

We first placed all three cohorts on the same context scale. The pooled reference contained 325,652 unique eligible people, of whom 65,374 had enough information to calculate a score. Most scored participants had three of the five components (52,799; 80.8%); 11,939 (18.3%) had four and 636 (1.0%) had all five. Income, education, food security, activity, and area deprivation were observed for 67,668, 83,850, 13,035, 2,538, and 312,209 reference participants, respectively. Relative to the 325,652-person pooled reference, the respective numbers without an observed value were 257,984, 241,802, 312,617, 323,114, and 13,443. These component counts describe recorded availability before score construction.

Among eligible SBP, LDL, and A1c participants, respectively, missing values numbered 231,950, 142,028, and 103,373 for income; 214,014, 138,870, and 98,612 for education; 292,038, 156,167, and 119,472 for food security; 303,882, 159,038, and 123,196 for activity; and 12,502, 5,594, and 5,443 for ZIP-linked deprivation. The score required at least three observed components, as described above.

Baseline eGFR was missing for 156,102, 26,940, and 17,418 eligible participants, respectively. Among participants with a computable context score, the strictly preindex creatinine result and derived eGFR were missing for 51,798, 2,917, and 3,936; preindex smoking responses were missing for 3,107, 815, and 962. A further 545, 335, and 460 scored participants had preindex eGFR below 60 mL/min/1.73 m^2^. These exclusions reconcile the score-computable and analyzed counts in Table 1. The coded age, sex, Black and Hispanic indicators, Charlson score, prior-year visits, index biomarker and date, observation end, and MACE follow-up had no null values in the eligible cohorts; model-ready records had no null adjustment variables.

**Table 1.** Selection from eligible source cohorts to the primary analysis.

| Cohort | Eligible | Score observed | Analyzed | MACE | Median index year analyzed / other |
| --- | --- | --- | --- | --- | --- |
| SBP | 306,353 | 71,868 | 19,525 (6.4%) | 144 | 2020 / 2016 |
| LDL | 161,937 | 19,830 | 16,578 (10.2%) | 129 | 2022 / 2015 |
| A1c | 126,504 | 22,935 | 18,539 (14.7%) | 191 | 2022 / 2017 |

*Eligible is the source cohort before the score and final renal-function restrictions. Score observed is the number with a computable preindex context score before the final renal-function filter. Analyzed is the final model population; percentages use Eligible as the denominator. MACE counts first postindex myocardial infarction, ischemic stroke, or heart failure in Analyzed. Median index year compares Analyzed with other eligible participants. SBP, systolic blood pressure; LDL, low-density lipoprotein cholesterol; A1c, hemoglobin A1c. Cohorts overlap.*

Requiring a computable score and the other model inputs narrowed each source cohort substantially (Table 1). Analyzed participants had later index dates than other eligible participants: the standardized difference in index year was 1.234 for SBP, 1.668 for LDL, and 1.446 for A1c. Their crude event rates were 2.88 versus 4.92, 3.52 versus 5.38, and 4.99 versus 6.83 events per 1,000 person-years, respectively. In the analyzed SBP, LDL, and A1c cohorts, mean ages were 40.7, 41.2, and 42.0 years; 65.5%, 68.1%, and 68.0% were female; and mean age-adjusted Charlson scores were 0.86, 0.94, and 1.03, respectively. Mean measured index values were 120.27 mmHg, 107.44 mg/dL, and 5.57%, respectively. Total observed follow-up was 50,046, 36,676, and 38,268 person-years (mean 2.56, 2.21, and 2.06 years per person), respectively. Recorded cohort-relevant medication eras active at index or ending within the preceding 180 days occurred in 20.1% of analyzed SBP, 6.4% of analyzed LDL, and 7.0% of analyzed A1c participants.

### Social context and subsequent events

At each cohort’s mean index biomarker value, a one-standard-deviation more favorable preindex score corresponded to adjusted MACE hazard ratios of 0.611 for SBP, 0.631 for LDL, and 0.544 for A1c (Table 2). Score-by-biomarker interaction p values were 0.4045, 0.5969, and 0.1766, respectively.

**Table 2.** Adjusted association of the fixed-reference social-context score with MACE.

| Cohort | Analyzed | MACE | Adjusted HR per SD (95% CI) | Score × biomarker p |
| --- | --- | --- | --- | --- |
| SBP | 19,525 | 144 | 0.611 (0.498–0.749) | 0.4045 |
| LDL | 16,578 | 129 | 0.631 (0.512–0.778) | 0.5969 |
| A1c | 18,539 | 191 | 0.544 (0.457–0.648) | 0.1766 |

*Adjusted hazard ratios compare a one-standard-deviation more favorable context score on the pooled reference scale, evaluated at each cohort's mean measured index biomarker value. Models include the measured biomarker, its square, the score-by-biomarker term, and the baseline covariates described in Methods. Score x biomarker p is the interaction term p value. HR, hazard ratio; CI, confidence interval; SD, standard deviation; MACE, major adverse cardiovascular events; SBP, systolic blood pressure; LDL, low-density lipoprotein cholesterol; A1c, hemoglobin A1c.*

Among participants with SBP below 140 mmHg, the event rate was 1.68 per 1,000 person-years in more favorable context and 3.39 in less favorable context. The corresponding rates were 2.05 versus 7.16 for LDL below 100 mg/dL and 2.34 versus 8.67 for A1c below 8% (Table 3). Within the LDL cohort, rates at or above 100 mg/dL were 1.52 versus 2.05 below that cutoff in more favorable context, and 6.60 versus 7.16 in less favorable context.

**Table 3.** Observed MACE rates by social context and measured biomarker category.

| Cohort | Context and measured biomarker | N | Events per 1,000 person-years |
| --- | --- | --- | --- |
| SBP | More favorable / below 140 | 8,983 | 1.68 |
| SBP | Both contexts / 140 or above | 2,181 | 5.76 |
| SBP | Less favorable / below 140 | 8,361 | 3.39 |
| LDL | More favorable / below 100 | 4,613 | 2.05 |
| LDL | More favorable / 100 or above | 6,342 | 1.52 |
| LDL | Less favorable / below 100 | 2,656 | 7.16 |
| LDL | Less favorable / 100 or above | 2,967 | 6.60 |
| A1c | More favorable / below 8 | 11,581 | 2.34 |
| A1c | Less favorable / below 8 | 6,392 | 8.67 |

*Context is split at the fixed pooled-reference median (−0.2573). Biomarker cutoffs are SBP <140 mmHg, LDL <100 mg/dL, and A1c <8%; the other category includes the cutoff. N is participants. Rates are unadjusted first postindex MACE events per 1,000 observed person-years. All four context-by-biomarker strata were calculated for each cohort. For display under the All of Us data dissemination policy, the two SBP at-or-above-cutoff rows are combined, the A1c at-or-above-cutoff rows are withheld, and the event-count column is omitted. This prevents direct or derived disclosure of event counts below the reporting threshold. These presentation changes did not alter cohort totals or model estimates. Cohorts overlap.*

### Restricted-era one-year sensitivity

Because analyzed participants were concentrated in later years, we repeated the analysis among those indexed in 2019–2023 and limited follow-up to one year. This included 13,943 SBP participants with 54 MACE events, 12,832 LDL participants with 62 events, and 14,073 A1c participants with 94 events. The adjusted one-year hazard ratios per standard deviation of more favorable context were 0.530 for SBP, 0.336 for LDL, and 0.365 for A1c (Table 4). A one-year event or at least one year of observation was recorded for 71.9%, 81.2%, and 80.8% of participants, respectively.

**Table 4.** One-year MACE association in the 2019–2023 index era.

| Cohort | Analyzed | One-year MACE | Adjusted HR per SD (95% CI) | One-year status known |
| --- | --- | --- | --- | --- |
| SBP | 13,943 | 54 | 0.530 (0.398–0.708) | 10,024 (71.9%) |
| LDL | 12,832 | 62 | 0.336 (0.263–0.428) | 10,414 (81.2%) |
| A1c | 14,073 | 94 | 0.365 (0.299–0.445) | 11,369 (80.8%) |

*Participants had index dates in 2019-2023; follow-up was capped at one year. The hazard ratio compares a one-standard-deviation more favorable context score on the pooled reference scale. Models stratify by index year and adjust for measured biomarker value and its square, age, sex, age-adjusted Charlson score, and preindex eGFR. One-year status known counts participants with MACE by one year or at least one year of recorded observation; earlier censoring contributed follow-up until observation ended. HR, hazard ratio; CI, confidence interval; SD, standard deviation; eGFR, estimated glomerular filtration rate; MACE, major adverse cardiovascular events.*

### Sensitivity analysis indexed at score availability

Moving the index to the first measurement after the score became computable produced SBP, LDL, and A1c cohorts of 54,338, 41,451, and 33,607 participants with 493, 288, and 316 events (Table 5). Each one-SD more favorable score was associated with lower hazard: HR 0.732 (95% CI 0.657–0.815), 0.660 (0.573–0.761), and 0.631 (0.552–0.722). Score-by-biomarker p values were 0.923, 0.322, and 0.052. Retaining participants without a preindex eGFR (123,248, 47,243, and 39,568 participants) gave HRs of 0.684 (0.636–0.736), 0.658 (0.575–0.753), and 0.647 (0.570–0.734).

**Table 5.**
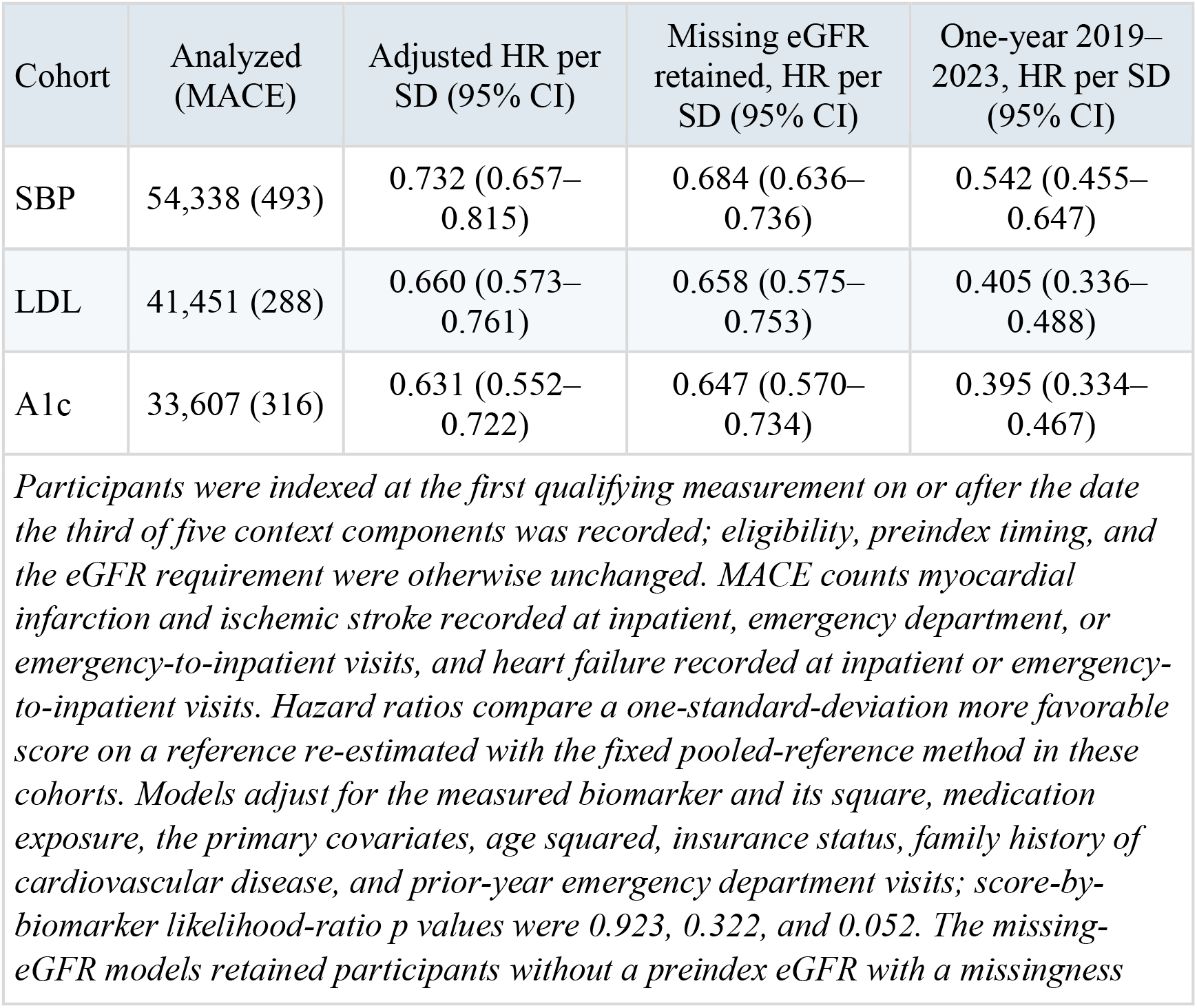
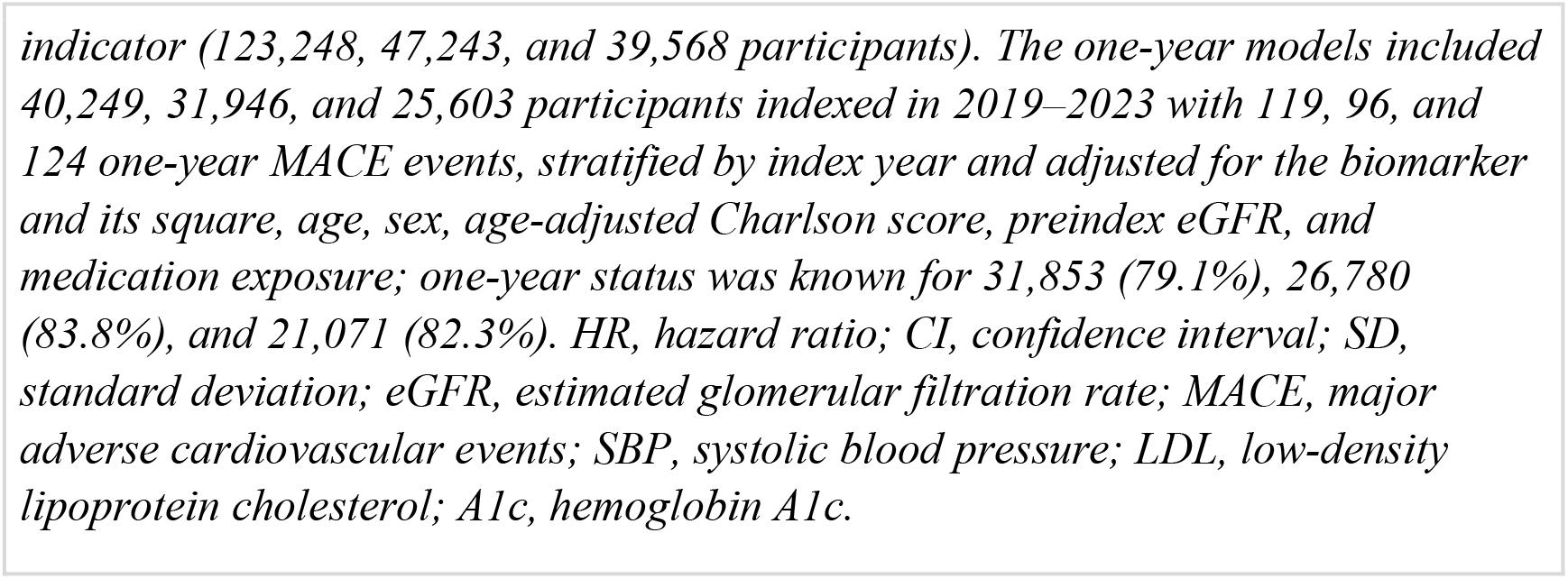
Sensitivity analysis indexed at score availability.

| Cohort | Analyzed (MACE) | Adjusted HR per SD (95% CI) | Missing eGFR retained, HR per SD (95% CI) | One-year 2019–2023, HR per SD (95% CI) |
| --- | --- | --- | --- | --- |
| SBP | 54,338 (493) | 0.732 (0.657–0.815) | 0.684 (0.636–0.736) | 0.542 (0.455–0.647) |
| LDL | 41,451 (288) | 0.660 (0.573–0.761) | 0.658 (0.575–0.753) | 0.405 (0.336–0.488) |
| A1c | 33,607 (316) | 0.631 (0.552–0.722) | 0.647 (0.570–0.734) | 0.395 (0.334–0.467) |

*Participants were indexed at the first qualifying measurement on or after the date the third of five context components was recorded; eligibility, preindex timing, and the eGFR requirement were otherwise unchanged. MACE counts myocardial infarction and ischemic stroke recorded at inpatient, emergency department, or emergency-to-inpatient visits, and heart failure recorded at inpatient or emergency-to-inpatient visits. Hazard ratios compare a one-standard-deviation more favorable score on a reference re-estimated with the fixed pooled-reference method in these cohorts. Models adjust for the measured biomarker and its square, medication exposure, the primary covariates, age squared, insurance status, family history of cardiovascular disease, and prior-year emergency department visits; score-by-biomarker likelihood-ratio p values were 0.923, 0.322, and 0.052. The missing-eGFR models retained participants without a preindex eGFR with a missingness*
*indicator (123,248, 47,243, and 39,568 participants). The one-year models included 40,249, 31,946, and 25,603 participants indexed in 2019–2023 with 119, 96, and 124 one-year MACE events, stratified by index year and adjusted for the biomarker and its square, age, sex, age-adjusted Charlson score, preindex eGFR, and medication exposure; one-year status was known for 31,853 (79.1%), 26,780 (83.8%), and 21,071 (82.3%). HR, hazard ratio; CI, confidence interval; SD, standard deviation; eGFR, estimated glomerular filtration rate; MACE, major adverse cardiovascular events; SBP, systolic blood pressure; LDL, low-density lipoprotein cholesterol; A1c, hemoglobin A1c.*

Below each cutoff, event rates in more versus less favorable context were 1.72 versus 3.86 per 1,000 person-years for SBP, 1.69 versus 5.47 for LDL, and 1.92 versus 5.97 for A1c. The corresponding adjusted HRs for less versus more favorable context were 1.55 (1.21–1.99), 2.01 (1.34–3.02), and 2.13 (1.57–2.89). The LDL pattern recurred. At or above 100 mg/dL, rates were 1.55 in more favorable and 3.93 in less favorable context, no higher than below the cutoff within either context group. Participants in less favorable context below 100 mg/dL had higher adjusted hazard than those in more favorable context at or above it (HR 2.26, 1.57–3.27). In the 2019–2023 one-year analysis of these cohorts, HRs were 0.542, 0.405, and 0.395.

## Discussion

Among participants with LDL cholesterol below 100 mg/dL, the recorded MACE rate was 2.05 per 1,000 person-years in the more favorable context group and 7.16 in the less favorable group. The same below-cutoff category therefore contained substantially different outcomes. The pattern also appeared below the SBP and A1c cutoffs. A favorable measurement remains useful information but it does not describe the whole person’s subsequent cardiovascular risk. The social-context score added information about who experienced an event after the models accounted for the measured biomarker itself.

These differences fit the pathway described by Powell-Wiley and colleagues, in which structural conditions and daily exposures shape biological responses, conventional risk factors, and cardiovascular outcomes (6). A clinic measurement captures one moment in that sequence. Income, education, and neighborhood deprivation belong beside SBP, LDL, and A1c in an outcome-focused account of risk. Prior studies linked individual and area-level social conditions to cardiovascular events (7–9). Our analysis locates that comparison within measured biomarker categories and finds variation in events among people below the same study cutoff.

The LDL cohort gives the clearest example of why a below-cutoff value cannot stand in for risk. Among participants below 100 mg/dL, the MACE rate was more than three times as high in less favorable as in more favorable context. Within either context group, the at-or-above-cutoff LDL rate was similar to or lower than the below-cutoff rate. A single achieved LDL value groups people with different clinical histories and medication exposure. In this cohort, recorded context distinguished subsequent events more clearly than the two LDL categories. A related ACCORD-BP analysis found that living arrangement modified the effect of combined intensive glycemic and blood-pressure treatment on cardiovascular events (19).

Target trials show why clinical outcomes and absolute effects belong beside changes in the biomarker. A synthesis of intensive glucose-lowering trials found clearer microvascular than macrovascular effects (20). In ACCORD, intensive glucose lowering increased all-cause mortality (HR 1.22) without significantly reducing the primary cardiovascular composite (21). Intensive blood-pressure treatment reduced stroke but not the primary composite and increased treatment-attributed serious adverse events (22); an individual-participant synthesis also documented benefit-harm tradeoffs (23). In primary prevention, statins reduced major vascular events (24), while a USPSTF synthesis estimated absolute reductions of 0.35 percentage points in all-cause mortality and 0.85 points in myocardial infarction (25). These endpoints, absolute effects, and harms are central to judging a target-focused strategy. Our cohort finding adds a practical fact: people below the same biomarker cutoff experienced different event rates according to recorded social context.

These findings carry the constraints of the records they come from, though they do align with broader available research. A computable score and the other required inputs were available for only 6.4% of SBP-eligible, 10.2% of LDL-eligible, and 14.7% of A1c-eligible participants, largely because most participants answered the surveys after their first qualifying measurement. Analyzed participants had later index dates and lower crude event rates than other eligible participants. When we instead indexed participants once their score became computable, the cohorts were 1.8 to 2.8 times larger. That analysis also restricted events to acute-care settings and used a broader adjustment set. The associations were somewhat smaller for SBP and A1c (HR 0.732 and 0.631 versus 0.611 and 0.544) and similar for LDL (0.660 versus 0.631), and the rate differences below each cutoff persisted. The score reflects the context that was recorded: 80.8% of computable reference scores used exactly three components, and food-security and activity responses were available for only 4.0% and 0.8% of reference participants. Its observed content was dominated by income, education, and area deprivation, with limited information on food security and activity. Selection into the analytic sample could still bias estimates in either direction, and replication in more completely characterized populations is needed.

Follow-up also affects the one-year sensitivity. In 2023, one-year outcome or observation support was 47.6% for SBP, 60.5% for LDL, and 62.4% for A1c. The Cox models used each participant’s observed time and depend on independent censoring within the modeled strata. Events above the A1c cutoff were sparse, limiting comparisons across all four A1c strata. Some participants entered more than one biomarker cohort, so the cohort-specific estimates describe overlapping populations. As in any observational analysis, residual confounding by unmeasured factors, including treatment intensity and adherence, cannot be excluded, and the score should be read as a marker of risk rather than a demonstrated causal target.

## Conclusion

A blood pressure, LDL cholesterol, or A1c value below a study cutoff did not mean uniformly low cardiovascular risk. Across three cohorts, recorded social context separated subsequent events after adjustment for the measured value and clinical factors. The association persisted in a recent-era one-year analysis and in larger cohorts indexed once the score became computable. Prevention programs measure success by the number, yet patients experience the event. Risk assessment that stops at the number is incomplete by design.

## Acknowledgments

The All of Us Research Program is supported by the National Institutes of Health, Office of the Director: Regional Medical Centers (OT2 OD026549, OT2 OD026554, OT2 OD026557, OT2 OD026556, OT2 OD026550, OT2 OD026552, OT2 OD026553, OT2 OD026548, OT2 OD026551, OT2 OD026555); Federally Qualified Health Centers (HHSN 263201600085U); Data and Research Center (5 U2C OD023196); Biobank (1 U24 OD023121); the Participant Center (U24 OD023176); Participant Technology Systems Center (1 U24 OD023163); Communications and Engagement (3 OT2 OD023205, 3 OT2 OD023206); and Community Partners (1 OT2 OD025277, 3 OT2 OD025315, 1 OT2 OD025337, 1 OT2 OD025276). The All of Us Research Program would not be possible without the partnership of its participants. The content is solely the responsibility of the authors and does not necessarily represent the official views of the National Institutes of Health.

## Disclosure

The authors report no conflicts of interest in this work.

**AI Acknowledgement**

The authors used generative AI large language models (OpenAI Codex and Anthropic Claude) to assist in coding and to reorganize the manuscript to journal format. Neither tool accessed participant-level data or performed any analyses; all analyses were run by the authors within the All of Us Researcher Workbench. The authors take full responsibility for the content.

## Ethics Approval and Informed Consent

The All of Us Institutional Review Board approved the Controlled Tier data resource (protocol 2021-02-TN-001). All participants provided informed consent for participation and authorization for research use of electronic health record data. This secondary analysis was conducted within the secure Researcher Workbench under the All of Us data-use agreement.

## Consent for Publication

Not applicable; the manuscript contains no identifiable individual information.

## Data Availability

Individual-level All of Us data cannot be publicly redistributed under the program’s data-use policy. Eligible researchers may apply for access through the All of Us Researcher Workbench (https://www.researchallofus.org/). The study workspace and analytic code can be made available to authorized All of Us researchers by the corresponding author, subject to program policy.

## Author Contributions

Conceptualization, data curation, and analysis were performed by J.M. The investigation and original draft were prepared by S.M. and J.M.; both authors contributed to review and editing.

## Abbreviations

A1c: hemoglobin A1c
CI: confidence interval
eGFR: estimated glomerular filtration rate
HR: hazard ratio
LDL: low-density lipoprotein cholesterol
MACE: major adverse cardiovascular events
SBP: systolic blood pressure.

